# Performance of a fully automated plasma tau phosphorylated at threonine 217 immunoassay to reflect amyloid-beta burden in an unselected cohort representative of clinical practice

**DOI:** 10.1101/2025.11.07.25339760

**Authors:** Sayuri Hortsch, Annunziata Di Domenico, Niels Borlinghaus, David Caley, Laura Kaminioti-Dumont, Sara Bohn Jeppesen, Armand González-Escalante, Craig Ritchie, Kristian Steen Frederiksen, Marc Suárez-Calvet

## Abstract

**Background:** With the emergence of disease-modifying anti-amyloid-beta (Aβ) therapies for Alzheimer’s disease (AD), early and accurate quantitative measures of Aβ burden are critical. Blood-based biomarkers are a scalable and minimally invasive diagnostic solution; plasma tau phosphorylated at threonine 217 (pTau217) is a promising marker for Aβ pathology. The clinical performance of the prototype Elecsys^®^ Phospho-Tau (217P) Plasma immunoassay (Roche Diagnostics) to detect Aβ burden was investigated in an unselected cohort reflective of clinical practice.

**Methods:** Plasma was prospectively collected from participants aged 55 to 80 years with objective or subjective cognitive impairment under evaluation for AD. Participants were recruited at multiple clinical sites spanning primary and secondary care. Plasma pTau217 concentrations measured using the prototype pTau217 plasma immunoassay were compared with amyloid positron emission tomography centiloid-based classification at different cutoffs, with further analyses performed at centiloid cutoff 30.

**Outcomes:** Among 588 participants, plasma pTau217 demonstrated high concordance with centiloid-based classification at selected cutoffs. The discriminative ability of plasma pTau217 to detect Aβ pathology peaked at centiloid cutoff 32 (area under the curve=0.933). Subgroup analyses at centiloid cutoff 30 demonstrated good discrimination of Aβ positivity/negativity by clinical diagnosis, age, and sex. Substantially impaired kidney function was found to influence plasma pTau217 levels.

**Interpretation:** The prototype pTau217 plasma immunoassay showed high accuracy in reflecting Aβ burden among individuals presenting with cognitive complaints across diverse clinical settings. These findings support its potential implementation into routine clinical practice to aid early detection of AD, alongside standard clinical and neuropsychologic assessments.

## 1. Introduction

Alzheimer’s disease (AD) is a progressive neurodegenerative disorder characterized by the accumulation of amyloid-beta (Aβ) plaques and tau tangles, leading to cognitive and functional impairment [1]. Treatment advances have seen the development of disease-modifying therapies (DMTs) that target Aβ plaques, such as donanemab and lecanemab [2,3]; however, these treatments are most effective in early stages of the disease [4]. Consequently, there is an urgent need for cost-effective, accessible, and minimally-invasive methods to identify individuals who may benefit from treatment with DMTs.

Blood-based biomarkers (BBBMs) have emerged as a promising solution for early detection of the disease [5]; of which, plasma tau phosphorylated at threonine 217 (pTau217) has consistently demonstrated high accuracy in identifying Aβ pathology in the brain [6]. The accuracy of the pTau217 biomarker was reinforced by the results of a global Round Robin study and a head-to-head study, where plasma pTau217 assays outperformed all other plasma phosphorylated tau variants tested [7,8]. Consequently, plasma pTau217 immunoassays, including the prototype Elecsys^®^ Phospho-Tau (217P) Plasma immunoassay (Roche Diagnostics International Ltd, Rotkreuz, Switzerland), are undergoing rigorous clinical validation and evaluation by regulatory bodies [9,10].

The established reference standard for *in vivo* confirmation of Aβ pathology in the brain is amyloid positron emission tomography (PET) and the approved Aβ detection method for clinical use is qualitative assessment through PET visual read. In recent years, the centiloid-based scale has been recommended by the Amyloid Imaging to Prevent Alzheimer’s Disease (AMYPAD) consortium for use within research settings and clinical trials for quantifying Aβ burden [11]. This scale expresses Aβ load via the measurement of a particular tracer (where 0 corresponds to tracer uptake in controls and 100 corresponds to average uptake in people with AD), which is harmonized across Aβ tracers and treatment centers [11,12]. Use of centiloid-based classification can depict the transition from the absence of pathology to subtle pathology and onto the presence of established pathology [12,13]. The cutoffs for centiloid-based classification are commonly selected to increase with disease severity, and their choice can therefore vary based on the research question and patient population. Centiloid cutoffs 24–30 generally represent the transition from Aβ positivity to Aβ negativity [12], cutoffs >30 reflect Aβ positivity with high certainty [11] and cutoffs 40–50 are reflective of a level of Aβ pathology associated with tau burden [12]. These centiloid-based classification cutoffs have not yet been validated for clinical diagnosis but offer an insightful measure of AD pathology [14] and, therefore, have been used in clinical trials of new Aβ-modifying therapies [15].

The primary objective of this study was to assess the clinical performance of the prototype Elecsys pTau217 plasma immunoassay versus different centiloid-based classification cutoffs to define possible corresponding plasma pTau217 cutoffs. While this analysis has been performed with other plasma pTau217 assays in clinical trial cohorts, to the best of our knowledge, this is the first time that such an analysis has been performed in an unselected cohort reflective of clinical practice, including individuals presenting with symptoms of cognitive decline across both primary and secondary care settings.

## 2. Methods

### 2.1. Study design and population

Plasma samples (K2-ethylenediaminetetraacetic acid) were collected from a prospective, multicenter cohort (part of the APPROACH [RD006263] study), across 12 clinical sites representative of primary and secondary care in Europe and the United States of America between May 2023 and March 2024 (Table S1). Samples were taken from adults aged 55 to 80 years with subjective cognitive decline (SCD), mild cognitive impairment (MCI), or mild dementia being evaluated for AD or other causes of cognitive decline. As part of the study, amyloid PET and magnetic resonance imaging (MRI) scans were performed. Data to assess the study population demographics and possible comorbidities were collected. The study was conducted according to the principles founded in the Declaration of Helsinki. Participants who consented for future use of samples and data were included in the present analysis.

### 2.2. Plasma biomarker analysis

After collection, plasma samples were stored at –80°C prior to analysis. Plasma pTau217 concentrations were measured retrospectively in a single run in August 2024 using the fully automated, high-throughput prototype Elecsys Phospho-Tau (217P) Plasma immunoassay on a Cobas^®^ e 801 analyzer (Roche Diagnostics International Ltd, Rotkreuz, Switzerland; reagents manufactured at Roche Diagnostics GmbH, Penzberg, Germany) at the Research and Development laboratory of Roche Diagnostics GmbH (Penzberg, Germany).

The prototype assay used in this study is a newer version (developed in 2024) [7,16] than that reported in studies from 2023 [17,18], with improved analytical sensitivity (i.e., lower limit of quantification). The instrument output is absolute plasma biomarker concentration. In this study, the discriminative ability of the prototype plasma pTau217 immunoassay to detect Aβ pathology was assessed using continuous data for the biomarker measurements. Absolute plasma biomarker concentrations (i.e., pg/mL values) are subject to change due to the prototype status of the assay.

### 2.3. Amyloid PET and centiloid-based classification

Amyloid PET was performed using one of the three U.S. Food and Drug Administration (FDA)-approved PET tracers: [18F]-Florbetapir, [18F]-Flutemetamol, or [18F]-Florbetaben [19–21]. Centiloids were calculated using MRI segmentation. Aβ positivity was defined according to centiloid-based classification at various cutoffs [11,12].

### 2.4. Statistical analysis

Inclusion in the population for analysis required participants to have both valid centiloid results and valid plasma pTau217 measurements. Baseline characteristics were summarized using descriptive analysis. The distributions of plasma pTau217 concentrations, overall and by centiloid-based classifications, were visualized using boxplots. Receiver operating characteristic analysis was performed to evaluate the discriminative ability of plasma pTau217 against centiloid-based classifications. Centiloid-based classification cutoffs were varied between 15 and 50, in intervals of one, to determine the dependence of the area under the curve (AUC) on the selected cutoff; the AUC and associated confidence intervals (CIs) were derived.

Cumulative distribution analysis was used to illustrate the dependency of positive percent agreement (PPA) and negative percent agreement (NPA) on the plasma pTau217 cutoff, against centiloid-based classifications of 24.1, 30, 40, and 50. To provide the results in tabular form, PPA and NPA with two-sided 95 % CIs (Wilson score method) were determined for plasma pTau217 cutoffs between 0.250 pg/mL and 0.800 pg/mL, in intervals of 0.025 pg/mL. Plasma pTau217 values equal to or greater than the cutoff values were interpreted as positive, while values below the cutoff were classified as negative. Additionally, plasma pTau217 performance at Youden’s index was assessed with respect to centiloid-based classifications [22].

For the different centiloid-based classification endpoints, a double cutoff approach was evaluated, where the lower cutoff was set at a target PPA of 90 % and the upper cutoff was set at a target NPA of 90 %. The double cutoff approach classified plasma pTau217 values into positive, negative, or indeterminate. Contingency tables were developed to summarize plasma pTau217 classification and Aβ status, together with percentage distribution of plasma pTau217 results.

A centiloid cutoff 30 has been shown to provide reliable detection of Aβ pathology [11], and was therefore of further interest in the context of this study. Investigations into plasma pTau217 concentrations were conducted in subgroups according to specific demographics and characteristics of interest. Boxplots were generated to explore the distribution of plasma pTau217 concentrations with respect to centiloid cutoff 30 and according to clinical diagnosis (SCD; MCI; mild dementia), age group (55 to 64 years; 65 to 74 years; 75 to 80 years), sex, and chronic kidney disease (CKD) staging, based on estimated glomerular filtration rate (eGFR) status: G1+G2 (≥60 mL/min/1:73 m^2^, mildly decreased to normal or high function), G3a (45 to 59 mL/min/1:73 m^2^, mildly to moderately decreased function), G3b (30 to 44 mL/min/1:73 m^2^, moderately to severely decreased function) and G4+G5 (≤29 mL/min/1:73 m^2^, severely decreased function to kidney failure) [23].

## 3. Results

### 3.1. Baseline characteristics

In total, 604 people were recruited to the study and 588 were eligible for inclusion in the analysis (Table 1); 16 people were excluded due to lack of paired centiloid and pTau217 results or lack of consent for secondary use of samples or data. Of the eligible participants, 189 (32.1 %) were clinically diagnosed with SCD, 352 (59.9 %) with MCI, and 40 (6.80 %) with mild dementia; seven (1.19 %) did not have an available clinical diagnosis.

**Table 1.**
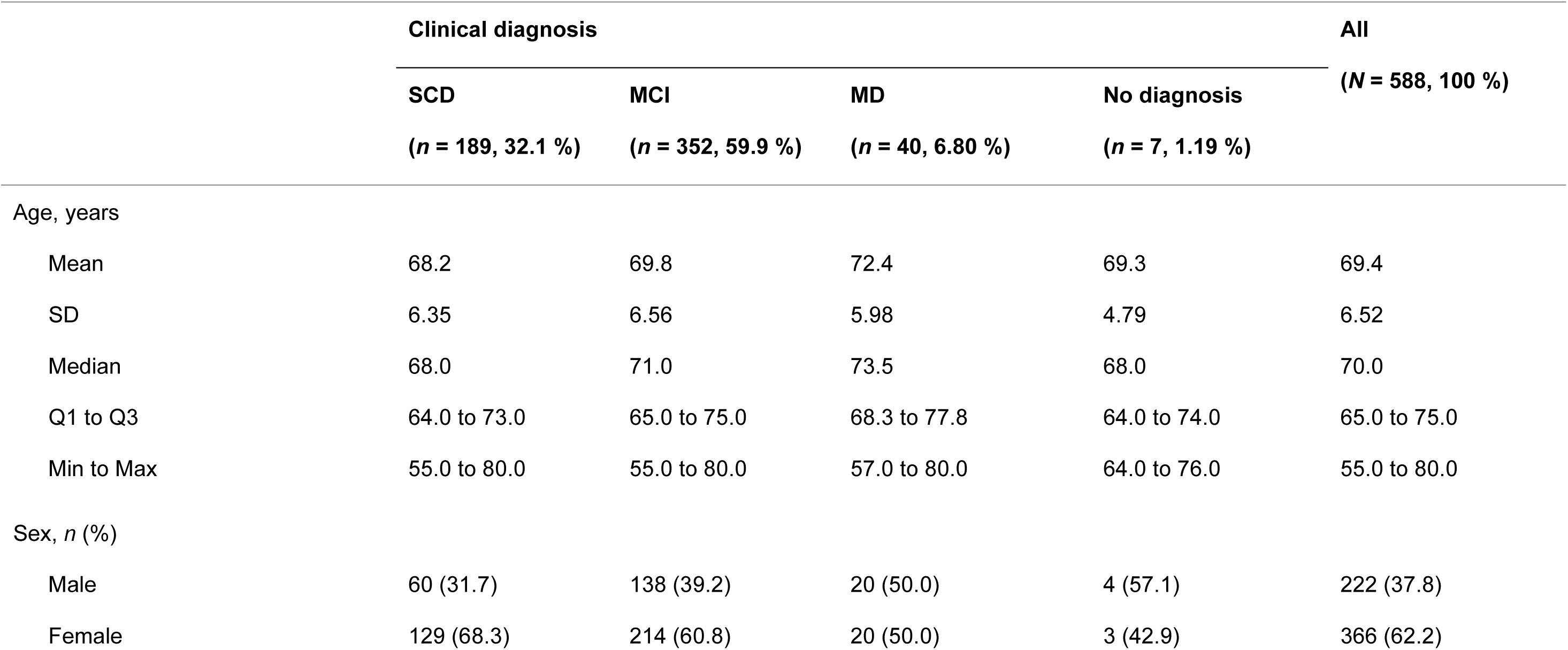
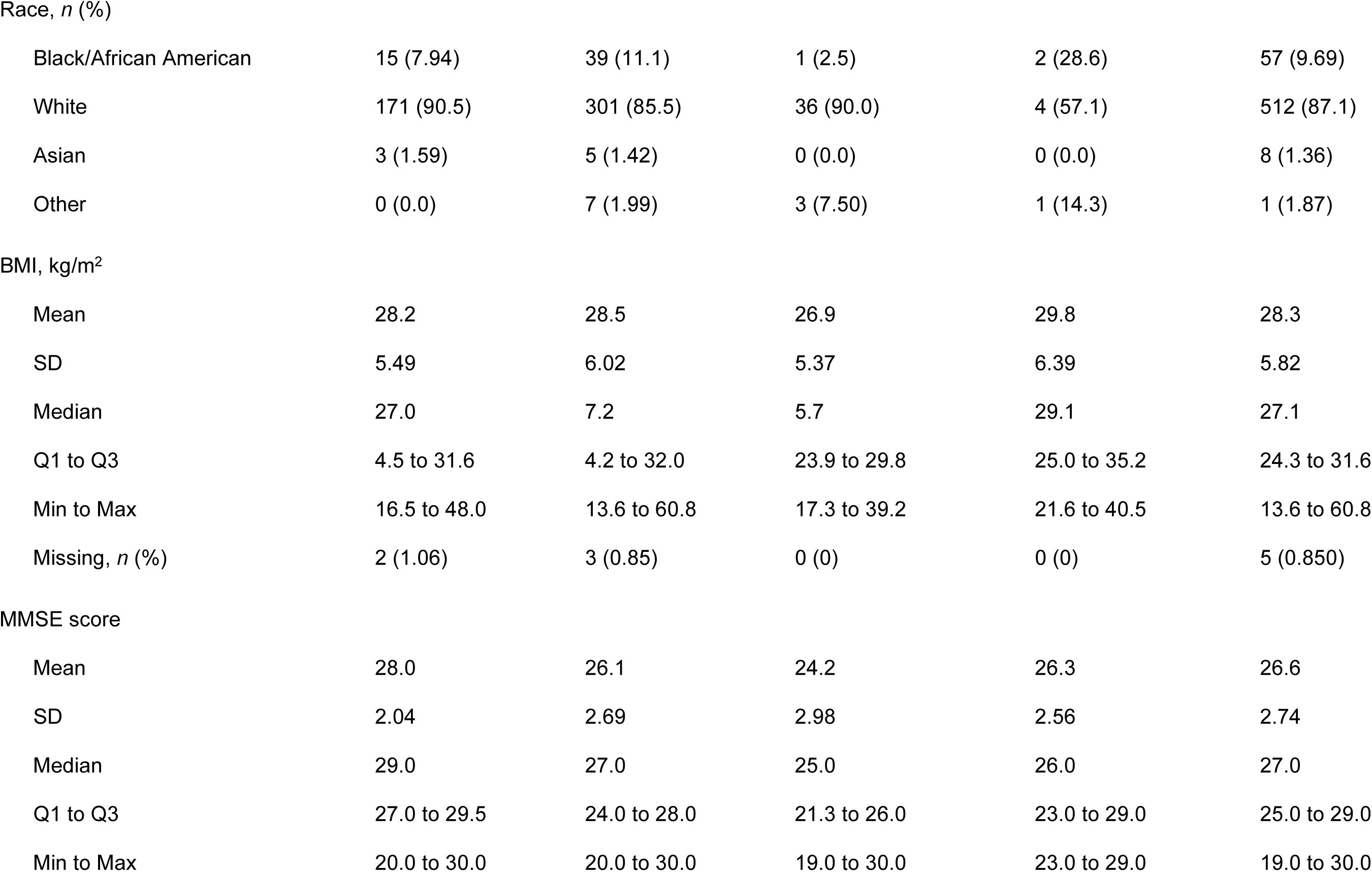
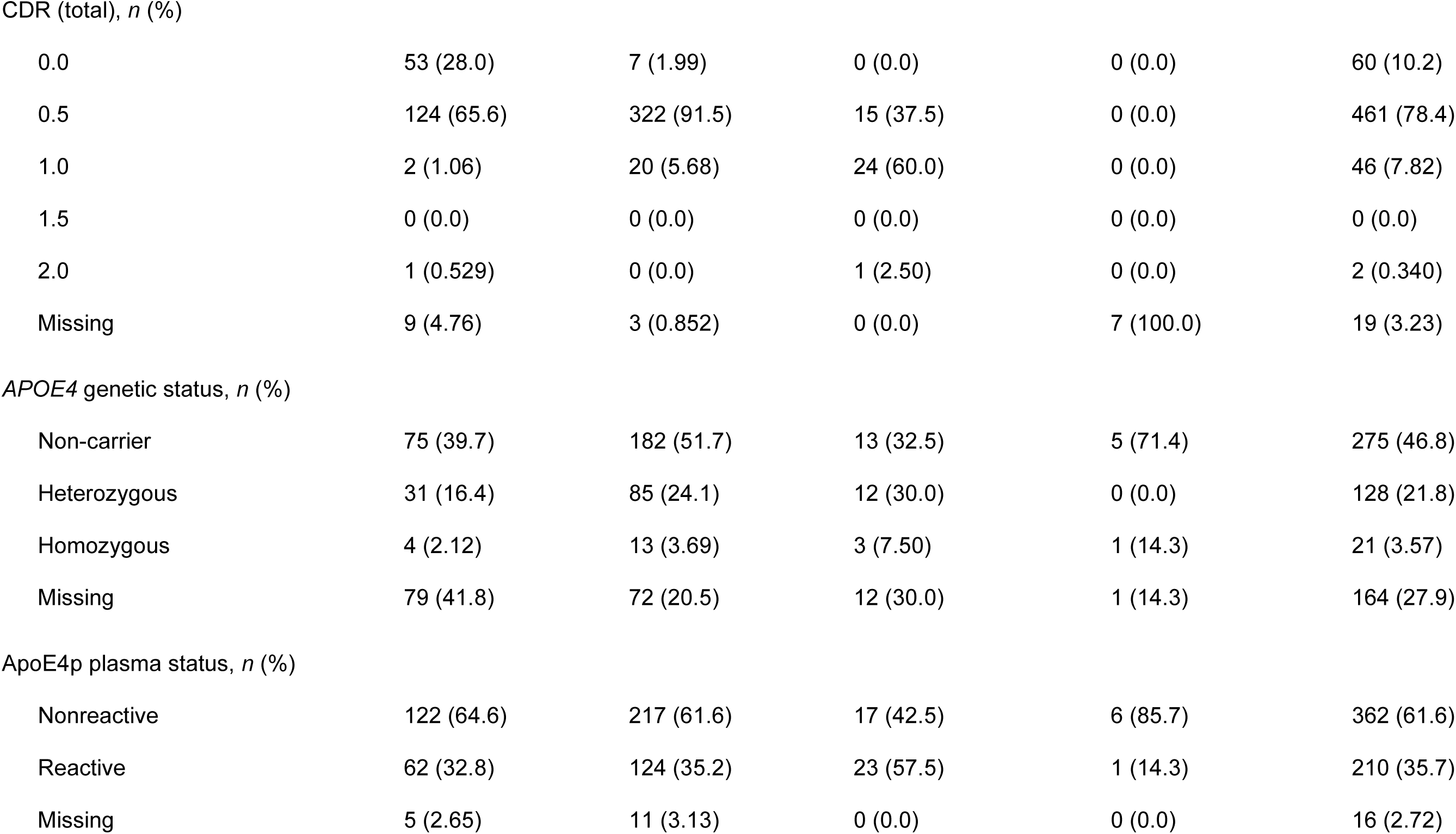
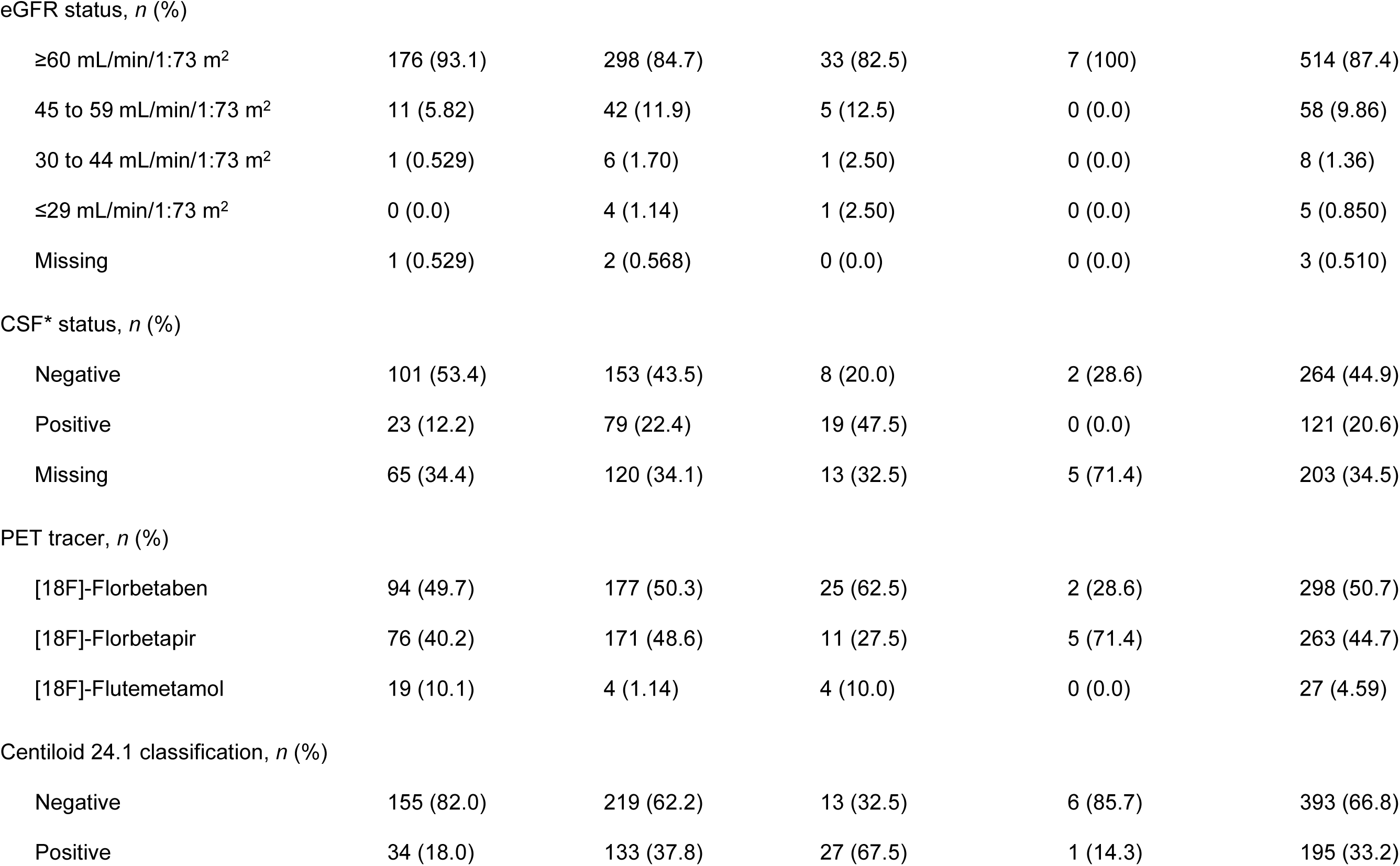
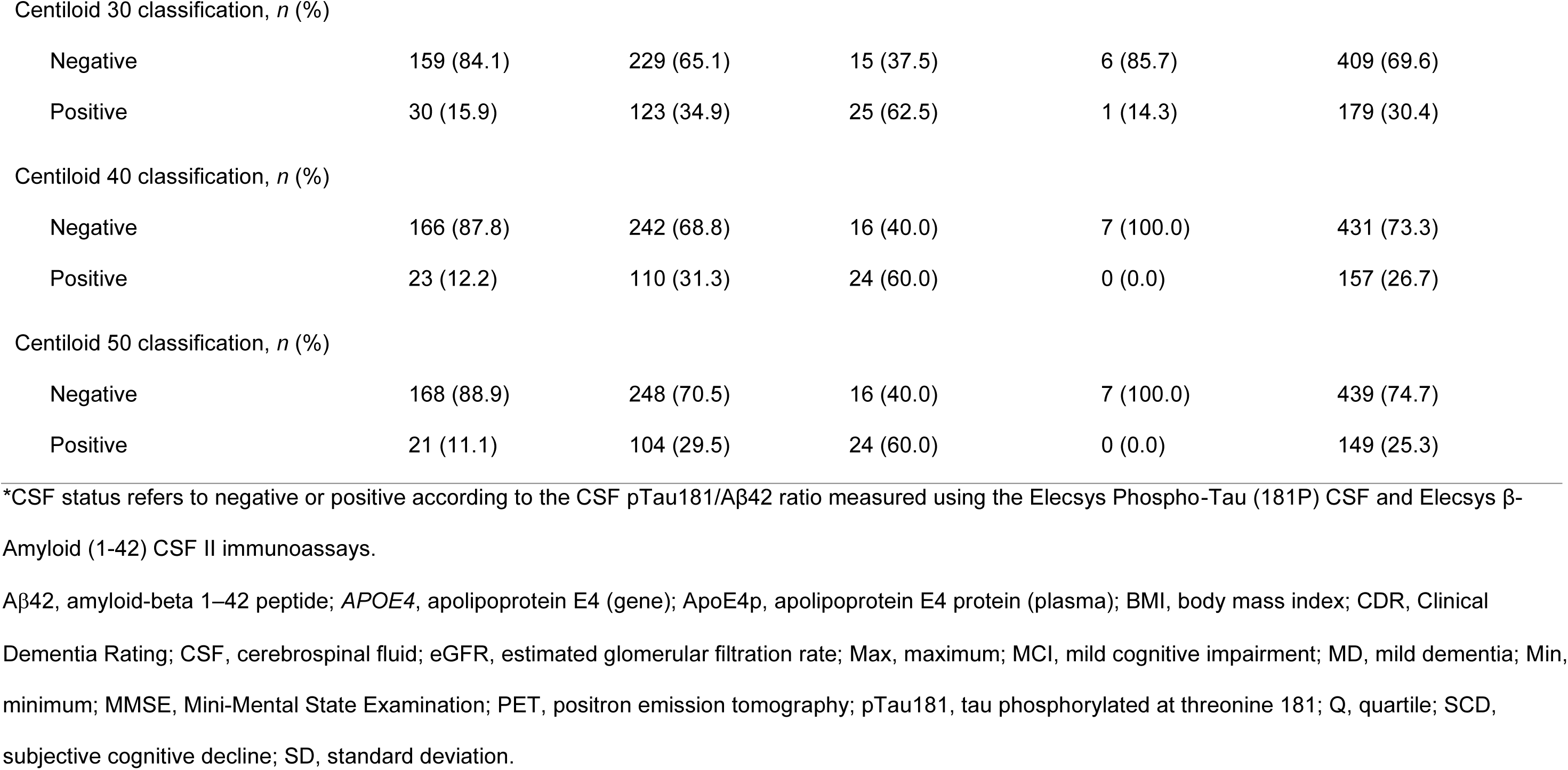
Baseline demographics and clinical characteristics (*N* = 588)

|  | Clinical diagnosis |  |  |  | All |
| --- | --- | --- | --- | --- | --- |
|  | SCD<br>(*n* = 189, 32.1 %) | MCI<br>(*n* = 352, 59.9 %) | MD<br>(*n* = 40, 6.80 %) | No diagnosis<br>(*n* = 7, 1.19 %) | (*N* = 588, 100 %) |
| Age, years |  |  |  |  |  |
| Mean | 68.2 | 69.8 | 72.4 | 69.3 | 69.4 |
| SD | 6.35 | 6.56 | 5.98 | 4.79 | 6.52 |
| Median | 68.0 | 71.0 | 73.5 | 68.0 | 70.0 |
| Q1 to Q3 | 64.0 to 73.0 | 65.0 to 75.0 | 68.3 to 77.8 | 64.0 to 74.0 | 65.0 to 75.0 |
| Min to Max | 55.0 to 80.0 | 55.0 to 80.0 | 57.0 to 80.0 | 64.0 to 76.0 | 55.0 to 80.0 |
| Sex, *n* (%) |  |  |  |  |  |
| Male | 60 (31.7) | 138 (39.2) | 20 (50.0) | 4 (57.1) | 222 (37.8) |
| Female | 129 (68.3) | 214 (60.8) | 20 (50.0) | 3 (42.9) | 366 (62.2) |
| Race, *n* (%) |  |  |  |  |  |
| Black/African American | 15 (7.94) | 39 (11.1) | 1 (2.5) | 2 (28.6) | 57 (9.69) |
| White | 171 (90.5) | 301 (85.5) | 36 (90.0) | 4 (57.1) | 512 (87.1) |
| Asian | 3 (1.59) | 5 (1.42) | 0 (0.0) | 0 (0.0) | 8 (1.36) |
| Other | 0 (0.0) | 7 (1.99) | 3 (7.50) | 1 (14.3) | 1 (1.87) |

BMI, kg/m<sup>2</sup>

|  |  |  |  |  |  |
| --- | --- | --- | --- | --- | --- |
| Mean | 28.2 | 28.5 | 26.9 | 29.8 | 28.3 |
| SD | 5.49 | 6.02 | 5.37 | 6.39 | 5.82 |
| Median | 27.0 | 7.2 | 5.7 | 29.1 | 27.1 |
| Q1 to Q3 | 4.5 to 31.6 | 4.2 to 32.0 | 23.9 to 29.8 | 25.0 to 35.2 | 24.3 to 31.6 |
| Min to Max | 16.5 to 48.0 | 13.6 to 60.8 | 17.3 to 39.2 | 21.6 to 40.5 | 13.6 to 60.8 |
| Missing, *n* (%) | 2 (1.06) | 3 (0.85) | 0 (0) | 0 (0) | 5 (0.850) |
| MMSE score |  |  |  |  |  |
| Mean | 28.0 | 26.1 | 24.2 | 26.3 | 26.6 |
| SD | 2.04 | 2.69 | 2.98 | 2.56 | 2.74 |
| Median | 29.0 | 27.0 | 25.0 | 26.0 | 27.0 |
| Q1 to Q3 | 27.0 to 29.5 | 24.0 to 28.0 | 21.3 to 26.0 | 23.0 to 29.0 | 25.0 to 29.0 |
| Min to Max | 20.0 to 30.0 | 20.0 to 30.0 | 19.0 to 30.0 | 23.0 to 29.0 | 19.0 to 30.0 |
| CDR (total), *n* (%) |  |  |  |  |  |
| 0.0 | 53 (28.0) | 7 (1.99) | 0 (0.0) | 0 (0.0) | 60 (10.2) |
| 0.5 | 124 (65.6) | 322 (91.5) | 15 (37.5) | 0 (0.0) | 461 (78.4) |
| 1.0 | 2 (1.06) | 20 (5.68) | 24 (60.0) | 0 (0.0) | 46 (7.82) |
| 1.5 | 0 (0.0) | 0 (0.0) | 0 (0.0) | 0 (0.0) | 0 (0.0) |
| 2.0 | 1 (0.529) | 0 (0.0) | 1 (2.50) | 0 (0.0) | 2 (0.340) |
| Missing | 9 (4.76) | 3 (0.852) | 0 (0.0) | 7 (100.0) | 19 (3.23) |
| *APOE4* genetic status, *n* (%) |  |  |  |  |  |
| Non-carrier | 75 (39.7) | 182 (51.7) | 13 (32.5) | 5 (71.4) | 275 (46.8) |
| Heterozygous | 31 (16.4) | 85 (24.1) | 12 (30.0) | 0 (0.0) | 128 (21.8) |
| Homozygous | 4 (2.12) | 13 (3.69) | 3 (7.50) | 1 (14.3) | 21 (3.57) |
| Missing | 79 (41.8) | 72 (20.5) | 12 (30.0) | 1 (14.3) | 164 (27.9) |
| ApoE4p plasma status, *n* (%) |  |  |  |  |  |
| Nonreactive | 122 (64.6) | 217 (61.6) | 17 (42.5) | 6 (85.7) | 362 (61.6) |
| Reactive | 62 (32.8) | 124 (35.2) | 23 (57.5) | 1 (14.3) | 210 (35.7) |
| Missing | 5 (2.65) | 11 (3.13) | 0 (0.0) | 0 (0.0) | 16 (2.72) |
| eGFR status, *n* (%) |  |  |  |  |  |
| ≥60 mL/min/1.73 m<sup>2</sup> | 176 (93.1) | 298 (84.7) | 33 (82.5) | 7 (100) | 514 (87.4) |
| 45 to 59 mL/min/1.73 m<sup>2</sup> | 11 (5.82) | 42 (11.9) | 5 (12.5) | 0 (0.0) | 58 (9.86) |
| 30 to 44 mL/min/1.73 m<sup>2</sup> | 1 (0.529) | 6 (1.70) | 1 (2.50) | 0 (0.0) | 8 (1.36) |
| ≤29 mL/min/1.73 m<sup>2</sup> | 0 (0.0) | 4 (1.14) | 1 (2.50) | 0 (0.0) | 5 (0.850) |
| Missing | 1 (0.529) | 2 (0.568) | 0 (0.0) | 0 (0.0) | 3 (0.510) |
| CSF* status, *n* (%) |  |  |  |  |  |
| Negative | 101 (53.4) | 153 (43.5) | 8 (20.0) | 2 (28.6) | 264 (44.9) |
| Positive | 23 (12.2) | 79 (22.4) | 19 (47.5) | 0 (0.0) | 121 (20.6) |
| Missing | 65 (34.4) | 120 (34.1) | 13 (32.5) | 5 (71.4) | 203 (34.5) |
| PET tracer, *n* (%) |  |  |  |  |  |
| [18F]-Florbetaben | 94 (49.7) | 177 (50.3) | 25 (62.5) | 2 (28.6) | 298 (50.7) |
| [18F]-Florbetapir | 76 (40.2) | 171 (48.6) | 11 (27.5) | 5 (71.4) | 263 (44.7) |
| [18F]-Flutemetamol | 19 (10.1) | 4 (1.14) | 4 (10.0) | 0 (0.0) | 27 (4.59) |
| Centiloid 24.1 classification, *n* (%) |  |  |  |  |  |
| Negative | 155 (82.0) | 219 (62.2) | 13 (32.5) | 6 (85.7) | 393 (66.8) |
| Positive | 34 (18.0) | 133 (37.8) | 27 (67.5) | 1 (14.3) | 195 (33.2) |
| Centiloid 30 classification, *n* (%) |  |  |  |  |  |
| Negative | 159 (84.1) | 229 (65.1) | 15 (37.5) | 6 (85.7) | 409 (69.6) |
| Positive | 30 (15.9) | 123 (34.9) | 25 (62.5) | 1 (14.3) | 179 (30.4) |
| Centiloid 40 classification, *n* (%) |  |  |  |  |  |
| Negative | 166 (87.8) | 242 (68.8) | 16 (40.0) | 7 (100.0) | 431 (73.3) |
| Positive | 23 (12.2) | 110 (31.3) | 24 (60.0) | 0 (0.0) | 157 (26.7) |
| Centiloid 50 classification, *n* (%) |  |  |  |  |  |
| Negative | 168 (88.9) | 248 (70.5) | 16 (40.0) | 7 (100.0) | 439 (74.7) |
| Positive | 21 (11.1) | 104 (29.5) | 24 (60.0) | 0 (0.0) | 149 (25.3) |

\*CSF status refers to negative or positive according to the CSF pTau181/Aβ 42 ratio measured using the Elecsys Phospho-Tau (181P) CSF and Elecsys β-Amyloid (1-42) CSF II immunoassays.
Aβ 42, amyloid-beta 1–42 peptide; *APOE4*, apolipoprotein E4 (gene); ApoE4p, apolipoprotein E4 protein (plasma); BMI, body mass index; CDR, Clinical Dementia Rating; CSF, cerebrospinal fluid; eGFR, estimated glomerular filtration rate; Max, maximum; MCI, mild cognitive impairment; MD, mild dementia; Min, minimum; MMSE, Mini-Mental State Examination; PET, positron emission tomography; pTau181, tau phosphorylated at threonine 181; Q, quartile; SCD, subjective cognitive decline; SD, standard deviation.

The study population was non-selective regarding sex, race, clinical disease stages, and comorbidities, reflective of clinical practice (Table 1). The mean age (standard deviation [SD]) in the overall study population was 69.4 (6.52) years and 62.2 % were female. Most of the study population were White (87.1 %); 9.69 % and 1.36 % of the participants were Black/African American and Asian, respectively. The mean (SD) body mass index according to clinical diagnosis was 28.2 (5.49) kg/m^2^ for participants with SCD, 28.5 (6.02) kg/m^2^ for participants with MCI, 26.9 (5.37) kg/m^2^ for participants with mild dementia, and 29.8 (6.39) kg/m^2^ for participants with an unknown clinical diagnosis. Regarding kidney function, 93.1 % of participants with SCD, 84.7 % of those with MCI, 82.5 % of those with mild dementia, and 100 % of those with an unknown clinical diagnosis had an eGFR status of G1+G2 (≥60 mL/min/1:73 m^2^, mildly decreased to normal or high function).

### 3.2. Distribution of plasma pTau217 concentrations and centiloid values

The mean (SD) plasma pTau217 concentration for the overall study population, measured using the prototype pTau217 plasma immunoassay, was 0.363 (0.333) pg/mL (Table 2). Only 17/588 (2.89 %) samples were below the lower end of the measuring range of the assay, highlighting the high analytical sensitivity of the assay. The mean (SD) plasma pTau217 concentrations were highest in participants with a clinical diagnosis of mild dementia (0.569 [0.321] pg/mL) compared with those with MCI (0.401 [0.375] pg/mL or SCD (0.252 [0.19] pg/mL) (Table 2; Fig. 1A) and participants without a clinical diagnosis (0.256 [0.188] pg/mL). Centiloid values also increased with more progressed disease stages (Fig. S1). A positive correlation was observed between plasma pTau217 concentrations and centiloid values (Pearson correlation coefficient: 0.623; Fig. 1B).

**Fig. 1.**
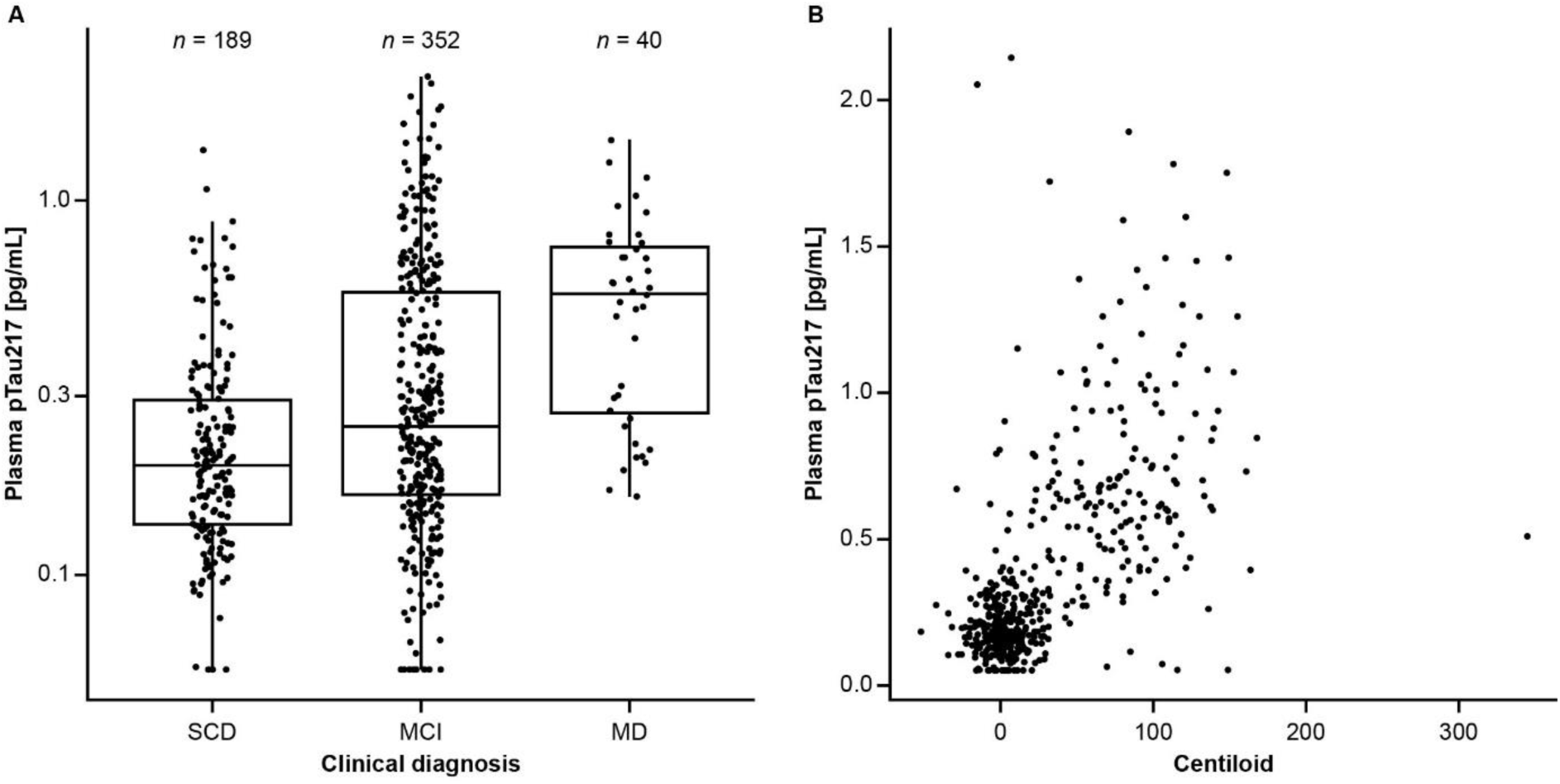
Distribution of plasma pTau217 concentrations with respect to A) clinical diagnosis and B) centiloid values. (A) shows a boxplot, with the box itself representing the interquartile range. The median is represented by a solid horizontal line within the box. The whiskers extend from the box to the most extreme values, excluding outliers. (B) shows a scatterplot of centiloid values versus plasma pTau217 results (Pearson correlation coefficient, restricted to plasma pTau217 values within measuring range: 0.623). MCI, mild cognitive impairment; MD, mild dementia; pTau217, tau phosphorylated at threonine 217; SCD, subjective cognitive decline.

**Table 2.** Distribution of plasma pTau217 overall and by clinical diagnosis.

|  | Clinical diagnosis |  |  |  | All |
| --- | --- | --- | --- | --- | --- |
|  | SCD<br>(*n* = 189, 32.1 %) | MCI<br>(*n* = 352, 59.9 %) | MD<br>(*n* = 40, 6.80 %) | No diagnosis<br>(*n* = 7, 1.19 %) | (*N* = 588, 100%) |
| Plasma pTau217, pg/mL |  |  |  |  |  |
| Mean | 0.252 | 0.401 | 0.569 | 0.256 | 0.363 |
| SD | 0.190 | 0.375 | 0.321 | 0.188 | 0.333 |
| Median | 0.196 | 0.250 | 0.564 | 0.189 | 0.226 |
| Q1 to Q3 | 0.136 to 0.295 | 0.163 to 0.571 | 0.265 to 0.762 | 0.104 to 0.442 | 0.157 to 0.463 |
| Min to Max | 0.0560 to 1.36 | 0.0560 to 2.14 | 0.162 to 1.45 | 0.0768 to 0.589 | 0.0560 to 2.14 |
| <LEoMR (*n*, %) | 3 (1.59) | 14 (3.98) | 0 (0.0) | 0 (0.0) | 17 (2.89) |
| >HEoMR (*n*, %) | 0 (0.0) | 0 (0.0) | 0 (0.0) | 0 (0.0) | 0 (0.0) |

HEoMR, higher end of measuring range; LEoMR, lower end of measuring range; Max, maximum; MCI, mild cognitive impairment; MD, mild dementia; Min, minimum; pTau217, tau phosphorylated at threonine 217; Q, quartile; SCD, subjective cognitive decline; SD, standard deviation.

### 3.3. Performance of plasma pTau217 with respect to centiloid-based classification

Depending on the chosen centiloid-based classification cutoff, Aβ positivity prevalence in the overall study population ranged from 33.2 % (centiloid cutoff 24.1) to 25.3 % (centiloid cutoff 50) (Table 1). Plasma pTau217 demonstrated high discriminative ability with respect to centiloid-based classification across a broad range of centiloid cutoffs (Fig. 2A; Fig. S2; Table S2), demonstrating that the prototype pTau217 plasma immunoassay can detect various degrees of Aβ burden. Screening of centiloid-based classification at all centiloid cutoffs between 15 and 50, in intervals of one, demonstrated that that the discriminative ability of plasma pTau217 peaked at centiloid cutoff 32, with an AUC of 0.933 (Table S2; Fig. S2). Notably, this centiloid cutoff is close to the centiloid cutoff ≥30 identified by the AMYPAD consortium as a threshold above which Aβ pathology can be detected with high certainty [11].

**Fig. 2.**
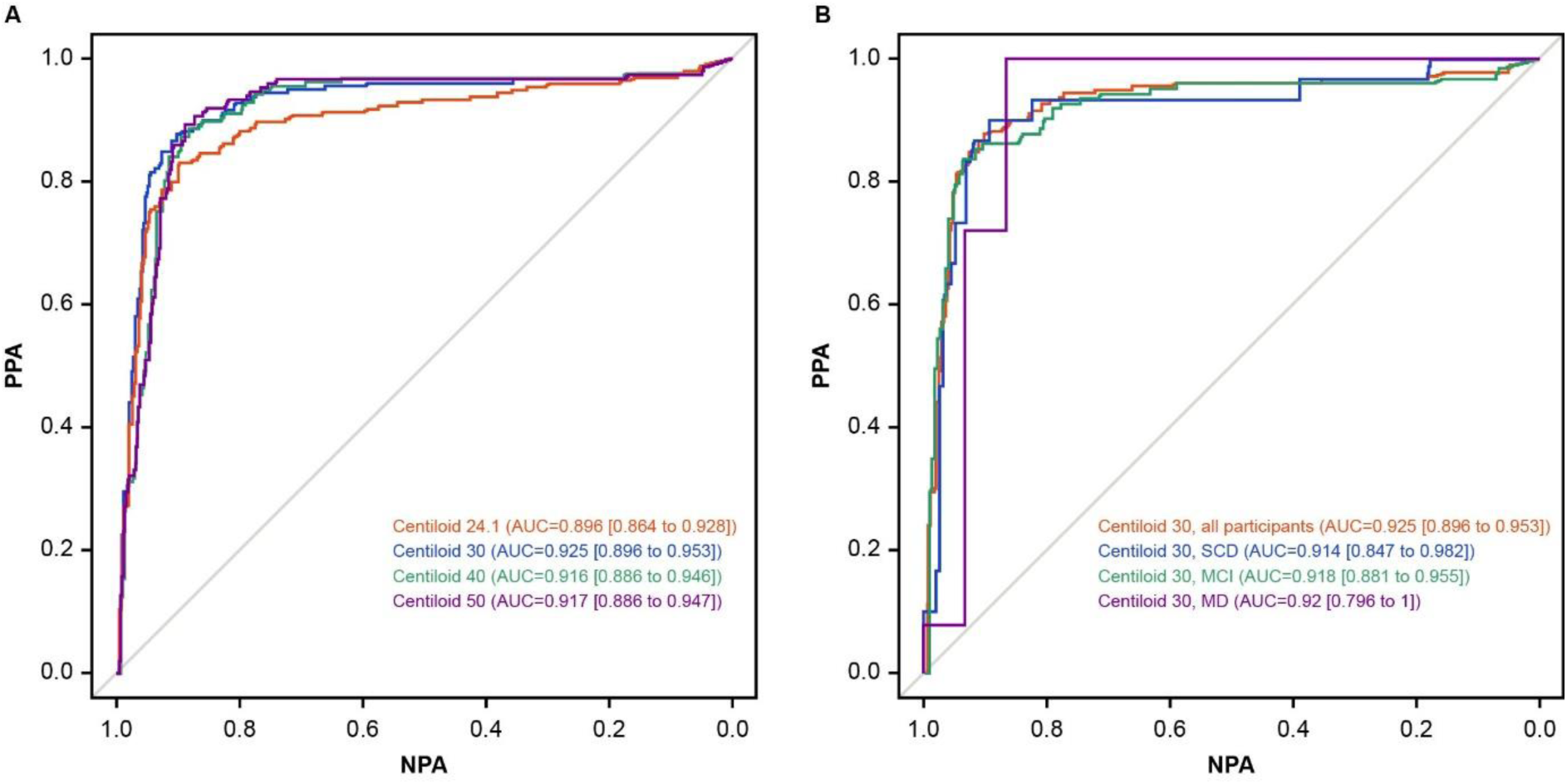
ROC analyses for plasma pTau217 by (A) centiloid-based classification at selected cutoffs and (B) clinical diagnosis at centiloid cutoff 30. AUC, area under the curve; MCI, mild cognitive impairment; MD, mild dementia; NPA, negative percent agreement; PPA, positive percent agreement; pTau217, tau phosphorylated at threonine 217; ROC, receiver operating characteristic; SCD, subjective cognitive decline.

The observed PPA and NPA values of the prototype pTau217 plasma immunoassay at a particular plasma pTau217 concentration varied according to the centiloid cutoff applied (Tables S3 to S6; Fig. S3). Table S7 presents the contingency of plasma pTau217 versus PET centiloid-based classification applying the centiloid 24.1, 30, 40, and 50 cutoffs, and when analyzed using a double cutoff approach (90 % PPA at the lower cutoff and 90 % NPA at the higher cutoff). All centiloid-based classifications resulted in less than 15 % of participants in the indeterminate zone, with only 5.78 % in the indeterminate zone for centiloid cutoff 30. These results show that, by selecting appropriate cutoffs for the prototype pTau217 plasma immunoassay, high concordance with the desired centiloid-based classification can be achieved.

### 3.4. Performance of plasma pTau217 with respect to centiloid-based classification cutoff 30 and the possible influence of comorbidities

The performance of the plasma pTau217 immunoassay at centiloid cutoff 30 was evaluated in different subgroups of interest to explore the potential impact of participant demographics/characteristics.

At centiloid cutoff 30, AUC values were 0.925 across all participants and were 0.914, 0.918, and 0.92 for participants with SCD, MCI, and mild dementia, respectively, demonstrating the consistently high discriminatory performance of plasma pTau217 (Fig. 2B).

Analysis of the distribution of plasma pTau217 concentrations applying a double cutoff approach (set at 90 % PPA and 90 % NPA) and with respect to centiloid cutoff 30 demonstrated good discrimination of Aβ positivity and negativity across all participants (Fig. 3A) and across subgroups of interest including by clinical diagnosis, age, and sex (Fig. 3B to 3D). As only 5.78 % of the tested participants presented with an indeterminate result (Table S7), 94.2 % of participants fell within the bounds (90 % certainty) for a positive or negative reading. Notably, participants with stage G3b to G5 CKD (eGFR ≤44 mL/min/1:73 m^2^, moderately decreased function to kidney failure), were found to have increased concentrations of plasma pTau217. This affected 13/588 (2.2 %) participants in the study (Fig. 3E). For participants with eGFR >30 mL/min/1:73 m^2^, the discriminative ability of plasma pTau217 was only minimally impacted; however, the few (<1 %; *n* = 5) participants with G4 to G5 CKD (≤29 mL/min/1:73 m^2^, severely decreased function to kidney failure) showed a considerable increase in plasma pTau217 concentrations.

**Fig. 3.**
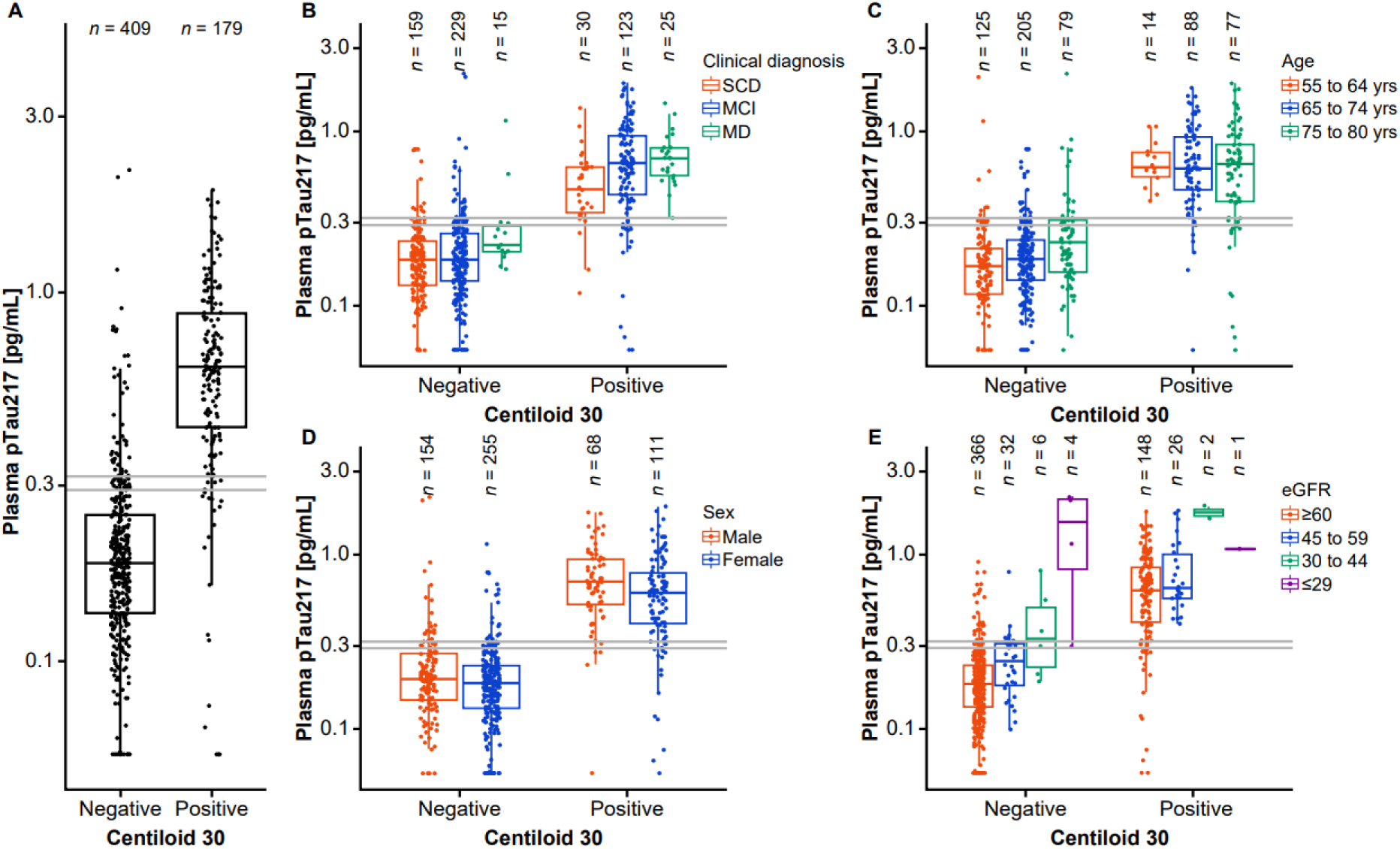
Distribution of plasma pTau217 concentrations with respect to centiloid cutoff 30 (A) across all participants and by (B) clinical diagnosis, (C) age, (D) sex, and (E) eGFR status, using a double cutoff approach set at 90 % PPA and 90 % NPA. The box itself represents the interquartile range. The median is represented by a solid horizontal line within the box. The whiskers extend from the box to the most extreme values, excluding outliers. The double cutoffs are represented by grey lines. eGFR status was determined according to KDIGO 2024 Clinical Practice Guideline for the Evaluation and Management of Chronic Kidney Disease (all values in mL/min/1:73 m^2^): ≥90, normal or high function (G1); 60 to 89, mildly decreased function (G2); 45 to 59, mildly to moderately decreased function (G3a); 30 to 44, moderately to severely decreased function (G3b); 15 to 29, severely decreased function (G4); and <15, kidney failure (G5). Thus, eGFR groupings for the purpose of analysis in this study were as follows (all values in mL/min/1:73 m^2^): G1+G2 (≥60, mildly decreased to normal or high function), G3a (45 to 59, mildly to moderately decreased function), G3b (30 to 44, moderately to severely decreased function) and G4+G5 (≤29, severely decreased function to kidney failure) [23]. eGFR, estimated glomerular filtration rate; G, group; KDIGO, Kidney Disease: Improving Global Outcomes; MCI, mild cognitive impairment; MD, mild dementia; NPA, negative percent agreement; PPA, positive percent agreement; pTau217, tau phosphorylated at threonine 217; SCD, subjective cognitive decline; yrs, years.

## 4. Discussion

In this study, we demonstrated that the prototype pTau217 plasma immunoassay was a highly accurate tool for detecting brain Aβ pathology in an unselected cohort reflective of clinical practice, in terms of sex, race, clinical disease stages (those in the early stages of cognitive impairment such as SCD, MCI, and mild dementia), and comorbidities. Plasma pTau217 was a good indicator of Aβ load, as concentrations increased with increasing centiloid values, and high concordance with centiloid values was observed across a range of centiloid cutoffs, demonstrating the ability of the biomarker to measure varying degrees of Aβ burden. The highest discriminative performance of plasma pTau217 was observed at centiloid cutoff 32; notably, this is in line with the centiloid cutoff ≥30 recommended by the AMYPAD Consortium as the threshold to define Aβ positivity with high certainty [11]. The findings of this study pave the way to address the urgent need for a cost-effective, globally accessible, and minimally-invasive method to aid in the early detection of individuals with AD, particularly as new DMTs become available [5,24].

As expected, the choice of centiloid-based classification cutoff value influenced the observed Aβ positivity prevalence, as well as the PPA and NPA values of the prototype pTau217 immunoassay at fixed concentrations, which should be considered when comparing the clinical performance of biomarker assays across studies. The discriminative ability of the pTau217 plasma immunoassay peaked at an AUC of 0.933 when using a quantitative centiloid cutoff of 32. A double cutoff approach for the plasma pTau217 immunoassay was explored with respect to centiloid-based classifications at cutoffs 24.1, 30, 40, and 50 to mitigate the ambiguity of borderline readings, which can affect 5 % to 20 % of individuals tested [25]. This method enhances clinical confidence by categorizing results as positive, negative, or indeterminate, with the indeterminate readings warranting further investigation [25]. For centiloid cutoff 30 specifically, a cutoff suggested by the AMYPAD Consortium for defining high-certainty Aβ positivity [11], the Elecsys pTau217 plasma immunoassay demonstrated excellent discriminatory performance, resulting in <6 % of individuals with plasma pTau217 measurements falling in the indeterminate zone, while the majority (>94 %) of measurements fell within the bounds for a positive or negative reading, with both false positive and false negative results being limited to a maximum of 10 %. The PPA and NPA values observed in this study align with Clinical Practice Guidelines, which recommend that BBBM tests demonstrating ≥90 % sensitivity and ≥75 % specificity may be used as triage tools, while those with ≥90 % sensitivity and specificity can replace amyloid PET imaging or cerebrospinal fluid (CSF) biomarker testing in individuals with cognitive impairment evaluated in specialized memory clinics [26]. While amyloid PET visual read is a qualitative approach that is the Conformité Européenne- and FDA-approved method for identifying Aβ pathology, the exploratory centiloid-based approach examined herein is quantitative and may be adjusted to the research question and study population [11,12].

A key finding of this study that is relevant for clinical practice is the influence of participant demographics and characteristics, including comorbidities, on plasma pTau217 concentrations. In agreement with previous studies [27,28], and explored specifically in the context of centiloid cutoff 30, this study found that plasma pTau217 concentrations were slightly increased with advanced age and CKD stage. Notably, increased plasma pTau217 concentrations were observed in participants with greatly impaired kidney function. In particular, Fig. 3E shows that several amyloid PET-negative participants with severe CKD had plasma pTau217 concentrations comparable to those seen in amyloid PET-positive participants (based on centiloid cutoff 30), meaning that these individuals could be wrongly classified as amyloid positive (false positives) based on plasma pTau217 levels alone.

This may be attributable to reduced renal clearance affecting pTau217 plasma concentrations, as well as other blood biomarker levels; however, it is important to highlight that this level of impairment (eGFR ≤29 mL/min/1:73 m^2^) was rare and affected <1 % of participants in this study. It is therefore recommended by clinical practice guidelines that BBBM tests (such as plasma pTau217 tests) should be interpreted with caution in individuals with severe CKD [26]. One potential avenue for further study could be to compare the use of other BBBMs with comparable renal function clearance to evaluate this effect.

The AUCs for discriminative performance of the pTau217 plasma immunoassay observed in this study were comparable to published data for other pTau217 assays [7,8,29–32], and to those reported for CSF biomarker tests [33,34]. Moreover, high correlation has been demonstrated between the prototype Elecsys pTau217 plasma immunoassay and the Lumipulse G pTau217 plasma immunoassay from Fujirebio [16]; in one of the reported head-to-head studies, the AUC for plasma pTau217 for detecting Aβ pathology was 0.907 (95 % CI: 0.858 to 0.957) with the prototype Elecsys pTau217 plasma immunoassay and 0.862 (95 % CI: 0.805 to 0.920) with the Lumipulse G pTau217 plasma immunoassay [16].

There is growing evidence that plasma pTau217 is a biomarker that can detect very early Aβ pathology [35]. A recent study determined plasma pTau217 levels in samples from cognitively impaired and unimpaired participants across five cohorts using the same prototype pTau217 plasma immunoassay as used in the present study. The high concordance with centiloid-based classification in cognitively impaired and unimpaired participants was confirmed in this study, as well as alterations in performance with different centiloid cutoffs. For cognitively impaired versus unimpaired participants, AUCs were 0.849 and 0.911, respectively, at centiloid cutoff 40, compared with AUCs of 0.872 and 0.864, respectively, at centiloid cutoff 24 (Hibar D, et al. manuscript in preparation). Future studies that assess performance with respect to centiloid-based classification should therefore consider the ability of selected cutoffs to detect Aβ pathology.

BBBM assays, such as the pTau217 plasma immunoassay, offer a solution to the drawbacks of current diagnostic standards. Amyloid PET scans are expensive, expose individuals to radiation, and have limited accessibility, particularly in remote areas due to the scarcity of scanners and specialized personnel. Similarly, lumbar punctures for CSF collection are invasive and painful for the individual [36]. In contrast, the Elecsys pTau217 plasma immunoassay requires a minimally invasive blood test and is fully automated with high-throughput and may offer advantages (e.g., speed, cost-effectiveness, and ability to process large numbers of samples) over other assays and methods that are in development (e.g., mass spectroscopy solutions).

The primary strength of this study was the unselected cohort, which is reflective of clinical practice. Participants with a spectrum of clinical disease stages and variety of comorbidities were included, in contrast to the highly selective participant populations typically enrolled in clinical trials. A limitation of this study was the use of a prototype assay with plasma samples measured in a single run at a single site, while sample collection was performed at multiple different sites. Absolute values of plasma pTau217 concentrations are subject to change and further validation is required, which is currently ongoing. Nonetheless, the prototype pTau217 plasma immunoassay represents a fully automated, high-throughput platform with the potential for global application to broaden access to AD diagnostics.

## 5. Conclusions

The prototype Elecsys pTau217 plasma immunoassay accurately detected Aβ positivity in an unselected cohort reflective of clinical practice, presenting with cognitive decline across primary and secondary care settings. The assay’s high accuracy and high throughput nature supports its implementation into routine clinical practice as a robust triage tool, which could aid in the early diagnosis of AD alongside neuropsychological assessments and other established clinical tests. By streamlining the AD diagnostic pathway and reducing the confirmatory testing bottleneck, this assay could improve identification of individuals with a high likelihood of Aβ pathology, thereby enabling timely access to DMTs.

## Ethics approval and consent to participate

This research was conducted in accordance with applicable ethical standards and regulatory guidelines. The study was conducted according to the International Council for Harmonisation Good Clinical Practice guidelines and the principles founded in the Declaration of Helsinki. The study protocol was approved by ethics committees/institutional review boards at the participating sites. Written informed consent for future use of samples and data was obtained from each participant prior to enrollment.

## Consent for publication

All authors involved in this research are aware of and agree with the content of the study and have consented to and contributed to the publication of the results.

## Declaration of competing interests

The authors declare the following financial interests/personal relationships which may be considered as potential competing interests:

Sayuri Hortsch reports a relationship with Roche Diagnostics GmbH that includes employment. Niels Borlinghaus reports a relationship with Roche Diagnostics GmbH that includes employment. Annunziata Di Domenico reports a relationship with Roche Diagnostics International Ltd that includes employment.

David Caley reports a relationship with Roche Diagnostics Ltd that includes employment. Laura Kaminioti-Dumont reports a relationship with Scottish Brain Sciences that includes employment.

Sara Bohn Jeppesen has no known competing financial interests or personal relationships that could have appeared to influence the work reported in this paper.

Armand González-Escalante has no known competing financial interests or personal relationships that could have appeared to influence the work reported in this paper.

Craig Ritchie reports a relationship with Scottish Brain Sciences that includes: employment (Chief Executive Officer, founder and majority shareholder of, which holds multiple commercial contracts including with Roche Pharmaceuticals, Roche Diagnostics, Thereni, MSD, Biogen, Lilly, Actinogen, Linus Healthcare, TauRx, and Kynexis), funding grants, and speaking and lecture fees.

Kristian Steen Frederiksen reports a relationship with Eisai, Novo Nordisk, Roche Diagnostics, and Eli Lilly (remuneration paid to institution) that includes: being a member of advisory boards or acts as consultant; also a relationship with Eisai/BioArctic, Eli Lilly, Novo Nordisk, and Roche Diagnostics (remuneration paid to institution) that includes: being engaged as a speaker; also reports a relationship with Biogen, Novo Nordisk, Roche, and Roche Diagnostics (remuneration paid to institution) that includes: being the principal investigator in clinical trials; also reports a relationship with Alzheimer’s Research and Therapy (Springer – Nature) that includes: Editor-in-Chief (personal remuneration); also reports a relationship with Aase og Ejner Danielsens Fond, Alzheimer Forskningsfonden, A.P. Møller Fonden, Beckett Fonden, C2N, DANMODIS, Ellen Mørch Fonden, ERA-PERMED, Fonden for Neurologisk Forskning, Grosserer F.L.Foghts Fond, Harboefonden, Hertzfonden, IHI, Innovationsfonden, Jascha Fonden, KID Fonden, Kong Christian den Tiendes Fond, Overretssagfører L. Zeuthens Mindelegat, Parkinsonforeningen, and Rigshospitalets Forskningspulje that includes: receiving research funding.

Marc Suárez-Calvet reports a relationship with Almirall, Eli Lilly, Quanterix, Novo Nordisk, and Roche Diagnostics that includes: receiving consultancy/speaker fees (paid to his institution); also reports a relationship with Eli Lilly, Grifols, Novo Nordisk, and Roche Diagnostics that includes: consultancy fees and served on advisory boards (paid to his institution); also reports a relationship with Roche Diagnostics that includes: granted a project and is a site investigator of a clinical trial (at his institution); also reports a relationship with ADx Neurosciences, Alamar Biosciences, ALZpath, Avid Radiopharmaceuticals, Eli Lilly, Fujirebio, Janssen Research & Development, Meso Scale Discovery, and Roche Diagnostics that includes: in-kind support for research (to his institution). Marc Suárez-Calvet did not receive any personal compensation from these organizations or any other for-profit organization.

## Credit authorship contribution statement

**Sayuri Hortsch:** Visualization, Writing – review & editing, Writing – original draft, Data curation, Resources, Formal analysis, Methodology, Conceptualization. **Annunziata Di Domenico:** Supervision, Visualization, Writing – review & editing, Writing – original draft, Data curation, Conceptualization. **Niels Borlinghaus:** Writing – review & editing, Resources, Validation, Methodology. **David Caley:** Project administration, Writing – review & editing, Resources, Investigation. **Laura Kaminioti-Dumont:** Writing – review & editing, Investigation. **Sara Bohn Jeppesen:** Writing – review & editing, Investigation. **Armand González-Escalante:** Writing – review & editing, Methodology. **Craig Ritchie:** Project administration, Supervision, Writing – review & editing, Writing – original draft, Investigation, Methodology, Conceptualization. **Kristian Steen Frederiksen:** Writing – review & editing, Investigation, Methodology. **Marc Suárez-Calvet:** Writing – review & editing, Methodology.

## Data availability

Data are available on reasonable request from the corresponding author of this publication.

## Acknowledgements

The authors would like to thank Alexander Jethwa (Roche Diagnostics GmbH, Penzberg, Germany) for his contributions to the conceptualization and methodology of the study and the reviewing and writing of the manuscript. Furthermore, the authors would like to thank Judith Schmitz (Chrestos GmbH, Essen, Germany) for her support in statistical analysis. COBAS and ELECSYS are trademarks of Roche. All other product names and trademarks are the property of their respective owners. The prototype Elecsys Phospho-Tau (217P) Plasma immunoassay is not approved for clinical use. The Elecsys Phospho-Tau (181P) CSF and Elecsys β-Amyloid (1-42) CSF II immunoassays are approved for clinical use.

