## Supplemental files for "Performance of a fully automated plasma tau phosphorylated at threonine 217 immunoassay to reflect amyloid-beta burden in an unselected cohort representative of clinical practice"

**Table S1.** Clinical study sites and number of samples ($N$ = 588) taken from each site

| Clinical site | Number of samples |
|---|---|
| Ace Alzheimer Center Barcelona, Spain | 83 |
| Adams Clinical, Watertown, MA, USA | 74 |
| Alzheimer's Research and Treatment Center, Wellington, FL, USA | 92 |
| Barcelonaβeta Brain Research Center, Barcelona, Spain | 18 |
| Charter Research Lady Lake, Lady Lake, FL, USA | 36 |
| Columbus Memory Center, Columbus, GA, USA | 75 |
| Danish Dementia Research Centre, Copenhagen, Denmark | 10 |
| Eastside Research Associates, Redmond, WA, USA | 34 |
| Genesis Neuroscience Clinic, Knoxville, TN, USA | 20 |
| K2 Medical Research Maitland, Maitland, FL, USA | 78 |
| K2 Medical Research, LLC Tampa, Tampa, FL, USA | 55 |
| Scottish Brain Sciences, Edinburgh, UK | 13 |

**Table S2.** ROC analyses for plasma pTau217 with respect to varying centiloid-based classification cutoffs

| Centiloid-based classification cutoff | AUC (95 % CI) |
|---|---|
| 15 | 0.85 (0.814 to 0.887) |
| 16 | 0.866 (0.832 to 0.9) |
| 17 | 0.873 (0.839 to 0.907) |
| 18 | 0.873 (0.839 to 0.907) |
| 19 | 0.883 (0.851 to 0.916) |
| 20 | 0.886 (0.853 to 0.919) |
| 21 | 0.891 (0.859 to 0.923) |
| 22 | 0.888 (0.856 to 0.921) |
| 23 | 0.891 (0.859 to 0.924) |
| 24 | 0.896 (0.864 to 0.928) |
| 25 | 0.896 (0.864 to 0.928) |
| 26 | 0.898 (0.866 to 0.93) |
| 27 | 0.907 (0.877 to 0.938) |
| 28 | 0.913 (0.883–0.943) |
| 29 | 0.917 (0.887 to 0.947) |
| 30 | 0.925 (0.896 to 0.953) |
| 31 | 0.929 (0.902 to 0.956) |
| 32 | 0.933 (0.906 to 0.96) |
| 33 | 0.931 (0.903 to 0.958) |
| 34 | 0.93 (0.903 to 0.958) |
| 35 | 0.927 (0.899 to 0.955) |

| | |
|---|---|
| 36 | 0.924 (0.896 to 0.953) |
| 37 | 0.924 (0.896 to 0.953) |
| 38 | 0.921 (0.892 to 0.95) |
| 39 | 0.919 (0.89 to 0.949) |
| 40 | 0.916 (0.886 to 0.946) |
| 41 | 0.916 (0.886 to 0.946) |
| 42 | 0.915 (0.885 to 0.945) |
| 43 | 0.917 (0.887 to 0.947) |
| 44 | 0.918 (0.888 to 0.948) |
| 45 | 0.916 (0.886 to 0.946) |
| 46 | 0.918 (0.888 to 0.948) |
| 47 | 0.918 (0.888 to 0.948) |
| 48 | 0.919 (0.889 to 0.949) |
| 49 | 0.917 (0.886 to 0.947) |
| 50 | 0.917 (0.886 to 0.947) |

The discriminative ability of pTau217 was reported in terms of AUC (CI) for each prespecified centiloid cutoff.

AUC, area under the curve; CI, confidence interval; pTau217, tau phosphorylated at threonine 217; ROC, receiver operating characteristic.

**Table S3.** Performance overview at various plasma pTau217 cutoff values with respect to centiloid 24.1

| Cutoff, pg/mL | PPA, % (95 % CI) | NPA, % (95 % CI) |
|---|---|---|
| 0.25 | 89.7 (84.7 to 93.3) | 77.1 (72.7 to 81.0) |
| 0.275 | 86.7 (81.2 to 90.7) | 81.2 (77.0 to 84.7) |
| 0.3 | 84.6 (78.9 to 89.0) | 85.8 (81.9 to 88.9) |
| **0.3165** | **83.1 (77.2 to 87.7)** | **89.8 (86.4 to 92.4)** |
| 0.325 | 80.0 (73.8 to 85.0) | 90.3 (87.0 to 92.9) |
| 0.35 | 78.5 (72.2 to 83.7) | 91.6 (88.4 to 94.0) |
| 0.375 | 75.9 (69.4 to 81.4) | 93.6 (90.8 to 95.7) |
| 0.4 | 72.8 (66.2 to 78.6) | 94.9 (92.3 to 96.7) |
| 0.425 | 70.8 (64.0 to 76.7) | 95.4 (92.9 to 97.1) |
| 0.45 | 67.2 (60.3 to 73.4) | 95.7 (92.3 to 97.3) |
| 0.475 | 64.6 (57.7 to 71.0) | 95.9 (93.5 to 97.5) |
| 0.5 | 63.1 (56.1 to 69.5) | 95.9 (93.5 to 97.5) |
| 0.525 | 61.0 (54.0 to 67.6) | 95.9 (93.5 to 97.5) |
| 0.55 | 57.9 (50.9 to 64.7) | 96.4 (94.1 to 97.9) |
| 0.575 | 54.9 (47.9 to 61.7) | 96.4 (94.1 to 97.9) |
| 0.6 | 52.3 (45.3 to 59.2) | 96.9 (94.7 to 98.2) |
| 0.625 | 46.2 (39.3 to 53.2) | 97.2 (95.1 to 98.4) |
| 0.65 | 43.1 (36.3 to 50.1) | 97.5 (95.4 to 98.6) |
| 0.675 | 40.0 (33.4 to 47.0) | 98.0 (96.0 to 99.0) |
| 0.7 | 35.9 (29.5 to 42.8) | 98.0 (96.0 to 99.0) |

| | | |
|---|---|---|
| 0.725 | 32.3 (26.1 to 39.2) | 98.0 (96.0 to 99.0) |
| 0.75 | 30.3 (24.2 to 37.0) | 98.0 (96.0 to 99.0) |
| 0.775 | 27.7 (21.9 to 34.4) | 98.0 (96.0 to 99.0) |
| 0.8 | 27.2 (21.4 to 33.8) | 98.7 (97.1 to 99.5) |

Youden's index and associated performance are shown in boldface.

CI, confidence interval; NPA, negative percent agreement; PPA, positive percent agreement; pTau217, tau phosphorylated at threonine 217.

**Table S4.** Performance overview at various plasma pTau217 cutoff values with respect to centiloid 30

| Cutoff, pg/mL | PPA,<br>% (95 % CI) | NPA,<br>% (95 % CI) |
|---|---|---|
| 0.25 | 94.4 (90.0 to 96.9) | 76.5 (72.2 to 80.4) |
| 0.275 | 92.2 (87.3 to 95.3) | 80.9 (76.8 to 84.4) |
| 0.3 | 89.9 (84.7 to 93.5) | 85.3 (81.6 to 88.4) |
| **0.319** | **87.7 (82.1 to 91.7)** | **90.2 (87.0 to 92.7)** |
| 0.325 | 86.6 (80.8 to 90.8) | 90.5 (87.2 to 92.9) |
| 0.35 | 84.9 (78.9 to 89.4) | 91.7 (88.6 to 94.0) |
| 0.375 | 82.1 (75.9 to 87.0) | 93.6 (90.8 to 95.6) |
| 0.4 | 78.8 (72.2 to 84.1) | 94.9 (92.3 to 96.6) |
| 0.425 | 76.5 (69.8 to 82.1) | 95.4 (92.9 to 97.0) |
| 0.45 | 72.6 (65.7 to 78.6) | 95.6 (93.2 to 97.2) |
| 0.475 | 69.8 (62.7 to 76.1) | 95.8 (93.4 to 97.4) |
| 0.5 | 68.2 (61.0 to 74.5) | 95.8 (93.4 to 97.4) |
| 0.525 | 65.9 (58.7 to 72.5) | 95.8 (93.4 to 97.4) |
| 0.55 | 62.6 (55.3 to 69.3) | 96.3 (94.0 to 97.8) |
| 0.575 | 59.8 (52.5 to 66.7) | 96.6 (94.3 to 98.0) |
| 0.6 | 57.0 (49.7 to 64.0) | 97.1 (94.9 to 98.3) |
| 0.625 | 50.3 (43.0 to 57.5) | 97.3 (95.2 to 98.5) |
| 0.65 | 46.9 (39.8 to 54.2) | 97.6 (95.6 to 98.7) |
| 0.675 | 43.6 (36.5 to 50.9) | 98.0 (96.2 to 99.0) |
| 0.7 | 39.1 (32.3 to 46.4) | 98.0 (96.2 to 99.0) |
| 0.725 | 35.2 (28.6 to 42.4) | 98.0 (96.2 to 99.0) |

| | | |
|---|---|---|
| 0.75 | 33.0 (26.5 to 40.1) | 98.0 (96.2 to 99.0) |
| 0.775 | 30.2 (23.9 to 37.3) | 98.0 (96.2 to 99.0) |
| 0.8 | 29.6 (23.4 to 36.7) | 98.8 (97.2 to 99.5) |

Youden’s index and associated performance are shown in boldface.

CI, confidence interval; NPA, negative percent agreement; PPA, positive percent agreement; pTau217, tau phosphorylated at threonine 217.

**Table S5.** Performance overview at various plasma pTau217 cutoff values with respect to centiloid 40

| Cutoff, pg/mL | PPA, % (95 % CI) | NPA, % (95 % CI) |
|---|---|---|
| 0.25 | 95.5 (91.1 to 97.8) | 73.3 (68.9 to 77.3) |
| 0.275 | 93.6 (88.7 to 96.5) | 77.7 (73.6 to 81.4) |
| 0.3 | 91.1 (85.6 to 94.6) | 81.9 (78.0 to 85.3) |
| 0.325 | 88.5 (82.6 to 92.6) | 87.2 (83.8 to 90.1) |
| 0.35 | 87.3 (81.1 to 91.6) | 88.6 (85.3 to 91.3) |
| **0.358** | **87.3 (81.1 to 91.6)** | **89.6 (86.3 to 92.1)** |
| 0.375 | 84.1 (77.5 to 89.0) | 90.5 (87.3 to 92.9) |
| 0.4 | 80.9 (74.0 to 86.3) | 91.9 (88.9 to 94.1) |
| 0.425 | 78.3 (71.3 to 84.1) | 92.3 (89.4 to 94.5) |
| 0.45 | 75.2 (67.9 to 81.3) | 93.0 (90.2 to 95.1) |
| 0.475 | 72.6 (65.2 to 79.0) | 93.5 (90.8 to 95.5) |
| 0.5 | 70.7 (63.2 to 77.3) | 93.5 (90.8 to 95.5) |
| 0.525 | 68.2 (60.5 to 74.9) | 93.5 (90.8 to 95.5) |
| 0.55 | 64.3 (56.6 to 71.4) | 94.0 (91.3 to 95.9) |
| 0.575 | 61.1 (53.3 to 68.4) | 94.2 (91.6 to 96.0) |
| 0.6 | 58.0 (50.1 to 65.4) | 94.7 (92.1 to 96.4) |
| 0.625 | 51.0 (43.2 to 58.7) | 95.1 (92.7 to 96.8) |
| 0.65 | 47.8 (40.1 to 55.5) | 95.6 (93.2 to 97.2) |
| 0.675 | 44.6 (37.0 to 52.4) | 96.3 (94.1 to 97.7) |
| 0.7 | 40.8 (33.4 to 48.6) | 96.8 (94.6 to 98.1) |
| 0.725 | 36.9 (29.8 to 44.7) | 97.0 (94.9 to 98.2) |

| | | |
|---|---|---|
| 0.75 | 34.4 (27.4 to 42.1) | 97.0 (94.9 to 98.2) |
| 0.775 | 31.8 (25.1 to 39.5) | 97.2 (95.2 to 98.4) |
| 0.8 | 31.2 (24.5 to 38.8) | 97.9 (96.1 to 98.9) |

Youden's index and associated performance are shown in boldface.

CI, confidence interval; NPA, negative percent agreement; PPA, positive percent agreement; pTau217, tau phosphorylated at threonine 217.

**Table S6.** Performance overview at various plasma pTau217 values with respect to centiloid 50

| **Cutoff, pg/mL** | **PPA,**<br>**% (95 % CI)** | **NPA,**<br>**% (95 % CI)** |
|---|---|---|
| 0.25 | 96.6 (92.4 to 98.6) | 72.4 (68.1 to 76.4) |
| 0.275 | 94.6 (89.8 to 97.3) | 76.8 (72.6 to 80.5) |
| 0.3 | 93.3 (88.1 to 96.3) | 81.3 (77.4 to 84.7) |
| 0.325 | 90.6 (84.8 to 94.3) | 86.6 (83.1 to 89.4) |
| 0.35 | 89.3 (83.3 to 93.3) | 87.9 (84.5 to 90.7) |
| **0.358** | **89.3 (83.3 to 93.3)** | **88.8 (85.5 to 91.5)** |
| 0.375 | 85.9 (79.4 to 90.6) | 89.7 (86.6 to 92.3) |
| 0.4 | 82.6 (75.7 to 87.8) | 91.1 (88.1 to 93.4) |
| 0.425 | 79.9 (72.7 to 85.5) | 91.6 (88.6 to 93.8) |
| 0.45 | 77.2 (69.8 to 83.2) | 92.5 (89.6 to 94.6) |
| 0.475 | 74.5 (66.9 to 80.8) | 92.9 (90.2 to 95.0) |
| 0.5 | 72.5 (64.8 to 79.0) | 92.9 (90.2 to 95.0) |
| 0.525 | 69.8 (62.0 to 76.6) | 92.9 (90.2 to 95.0) |
| 0.55 | 66.4 (58.5 to 73.5) | 93.6 (90.9 to 95.6) |
| 0.575 | 63.1 (55.1 to 70.4) | 93.8 (91.2 to 95.7) |
| 0.6 | 59.7 (51.7 to 67.3) | 94.3 (91.7 to 96.1) |
| 0.625 | 52.3 (44.4 to 60.2) | 94.8 (92.3 to 96.5) |
| 0.65 | 49.7 (41.7 to 57.6) | 95.4 (93.1 to 97.0) |
| 0.675 | 46.3 (38.5 to 54.3) | 96.1 (93.9 to 97.6) |
| 0.7 | 42.3 (34.6 to 50.3) | 96.6 (94.4 to 97.9) |
| 0.725 | 38.3 (30.8 to 46.3) | 96.8 (94.7 to 98.1) |

| | | |
|---|---|---|
| 0.75 | 35.6 (28.3 to 43.5) | 96.8 (94.7 to 98.1) |
| 0.775 | 32.9 (25.9 to 40.8) | 97.0 (95.0 to 98.3) |
| 0.8 | 32.2 (25.2 to 40.1) | 97.7 (95.9 to 98.8) |

Youden's index and associated performance are shown in boldface.

CI, confidence interval; NPA, negative percent agreement; PPA, positive percent agreement; pTau217, tau phosphorylated at threonine 217.

**Table S7.** Contingency table of plasma pTau217 versus PET centiloid-based classification at different cutoffs

| | | **Positive, *n* (%)** | **Negative, *n* (%)** | **Sum, *n* (%)** | **LR (95 % CI)** | **PV, % (95 % CI)** |
|---|---|---|---|---|---|---|
| Centiloid 24.1 status at 33.2 % prevalence; lower cutoff=0.234 pg/mL, upper cutoff=0.317 pg/mL | Positive | 160 (82.1) | 39 (9.92) | 199 (33.8) | 8.27 (6.09 to 11.2) | 80.4 (75.1 to 84.8) |
| | Indeterminate | 16 (8.21) | 69 (17.6) | 85 (14.5) | 0.467 (0.279 to 0.783) | 18.8 (12.2 to 28.0) |
| | Negative | 19 (9.74) | 285 (72.5) | 304 (51.7) | 0.134 (0.0873 to 0.207) | 6.25 (4.15 to 9.31) |
| | Total | 195 | 393 | 588 (100) | — | — |
| Centiloid 30 status at 30.4 % prevalence; lower cutoff=0.291 pg/mL, upper cutoff=0.318 pg/mL | Positive | 157 (87.7) | 40 (9.78) | 197 (33.5) | 8.97 (6.65 to 12.1) | 79.7 (74.4 to 84.1) |
| | Indeterminate | 5 (2.79) | 29 (7.09) | 34 (5.78) | 0.394 (0.155 to 1.00) | 14.7 (6.35 to 30.5) |
| | Negative | 17 (9.50) | 340 (83.1) | 357 (60.7) | 0.114 (0.0725 to 0.180) | 4.76 (3.08 to 7.30) |
| | Total | 179 | 409 | 588 (100) | — | — |
| Centiloid 40 status at 26.7 % prevalence; lower cutoff=0.305 pg/mL, upper cutoff=0.364 pg/mL | Positive | 133 (84.7) | 43 (9.98) | 176 (29.9) | 8.49 (6.35 to 11.4) | 75.6 (69.8 to 80.5) |
| | Indeterminate | 10 (6.37) | 33 (7.66) | 43 (7.31) | 0.832 (0.420 to 1.65) | 23.3 (13.3 to 37.5) |
| | Negative | 14 (8.92) | 355 (82.4) | 369 (62.8) | 0.108 (0.0655 to 0.179) | 3.79 (2.33 to 6.12) |
| | Total | 157 | 431 | 588 (100) | — | — |

| | | | | | | |
|---|---|---|---|---|---|---|
| Centiloid 50 status at 25.3 % prevalence; lower cutoff=0.337 pg/mL, upper cutoff=0.387 pg/mL | Positive | 128 (85.9) | 43 (9.79) | 171 (29.1) | 8.77 (6.55 to 11.7) | 74.9 (69.0 to 79.9) |
| | Indeterminate | 7 (4.70) | 12 (2.73) | 19 (3.23) | 1.72 (0.689 to 4.28) | 36.8 (19.0 to 59.3) |
| | Negative | 14 (9.40) | 384 (87.5) | 398 (67.7) | 0.107 (0.0652 to 0.177) | 3.52 (2.16 to 5.67) |
| | Total | 149 | 439 | 588 (100) | — | — |

CI, confidence interval; LR, likelihood ratio; PET, positron emission tomography; pTau217, tau phosphorylated at threonine 217; PV, predictive value.

**Fig. S1.** Centiloid distribution by clinical diagnosis.

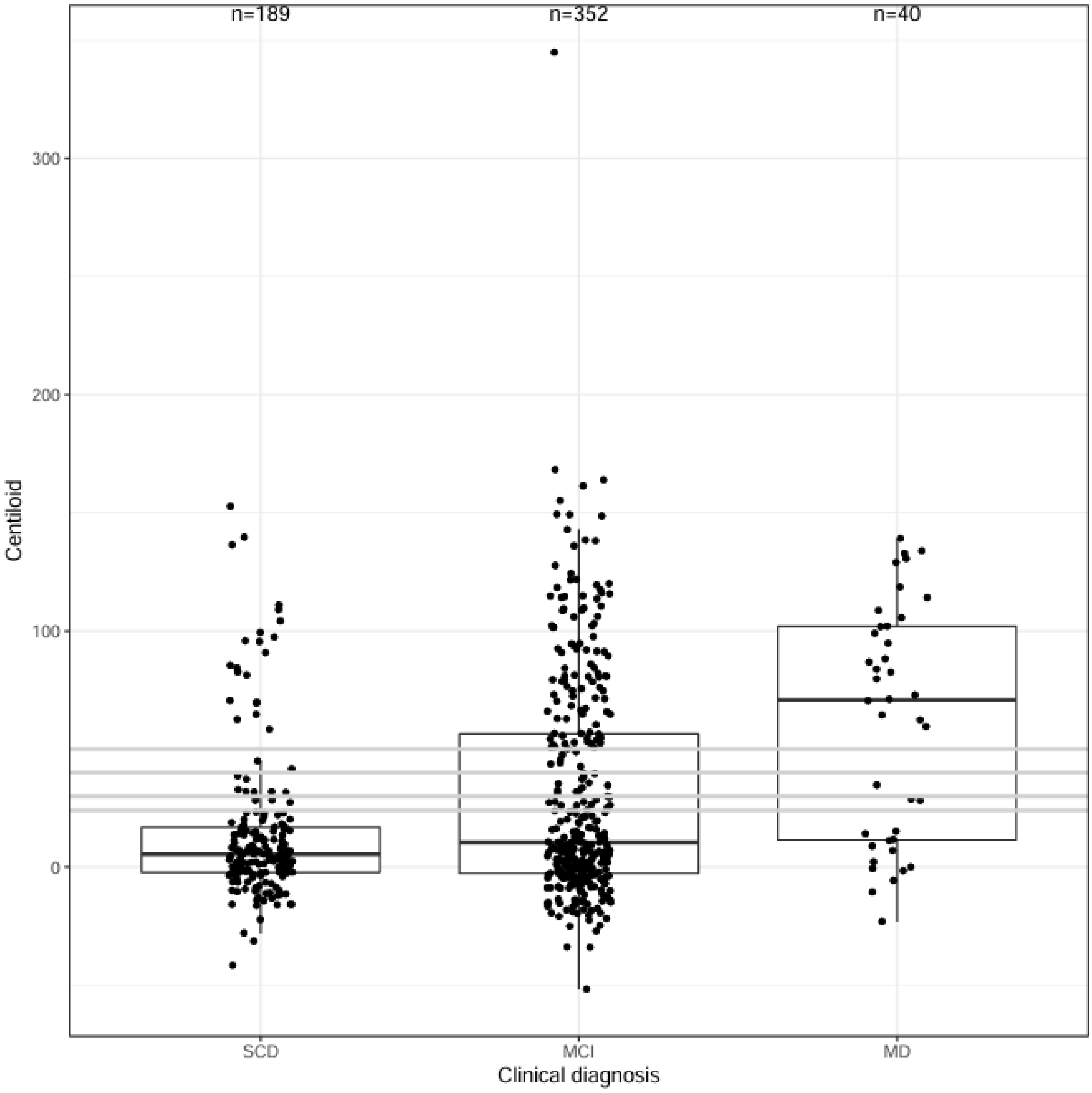


The applied centiloid cutoffs are shown by the grey lines; 24.1, 30, 40, and 50.

MCI, mild cognitive impairment; MD, mild dementia; SCD, subjective cognitive decline.

**Fig. S2**. ROC analyses for plasma pTau217 with respect to varying centiloid-based classification cutoffs.

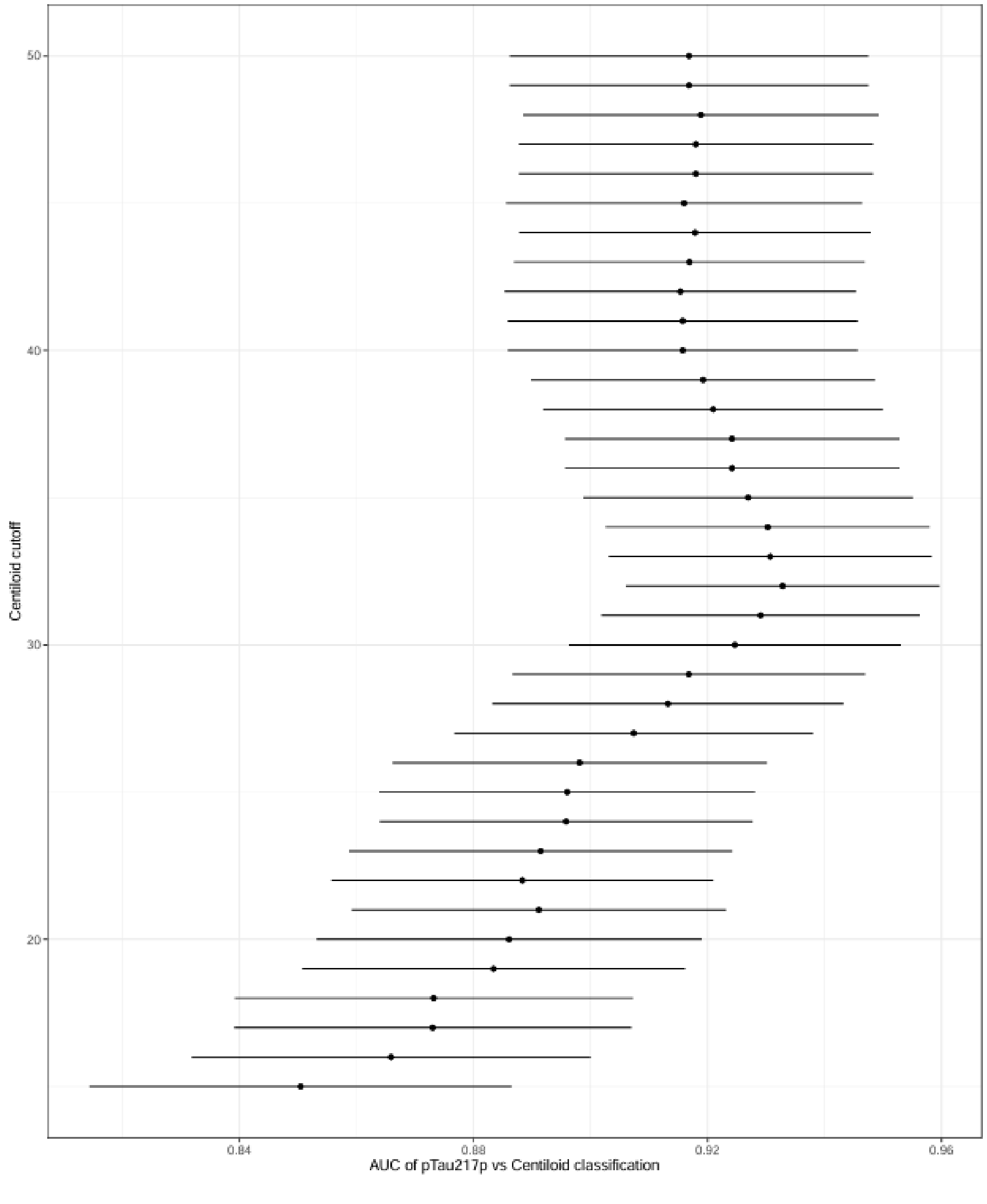


AUC, area under the curve; pTau217, tau phosphorylated at threonine 217; ROC, receiver operating characteristic.

**Fig. S3.** Cumulative distribution analysis illustrating the PPA and NPA of the prototype pTau217 plasma immunoassay with respect to centiloid-based classification at selected cutoffs.

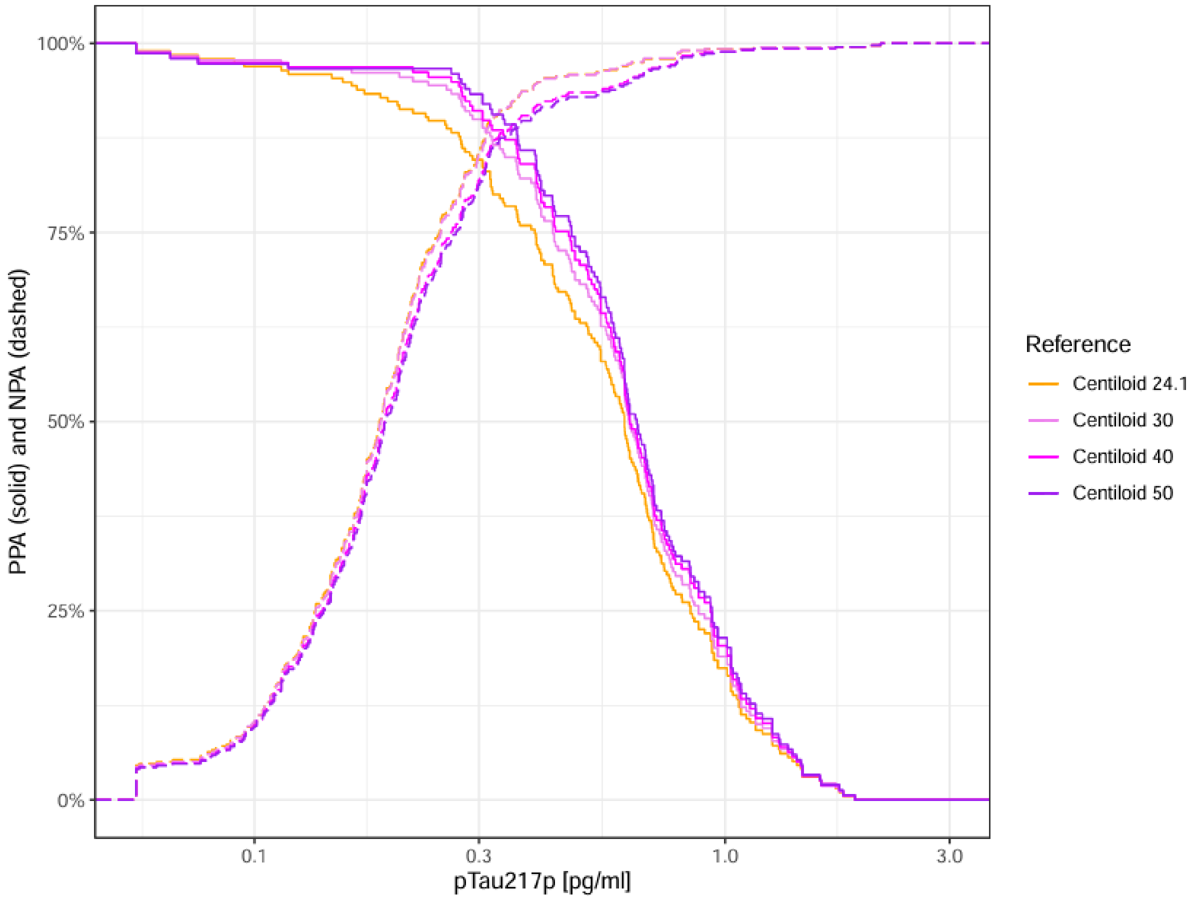


NPA, negative percent agreement; PPA, positive percent agreement; pTau217, tau phosphorylated at threonine 217.
